# Study of the Sudanese perceptions of COVID-19: Applying the Health Belief Model

**DOI:** 10.1101/2020.05.28.20115477

**Authors:** Elwalid F. Nasir, Hatim Mohammed Almahdi Yagoub, Ahmed Khalid Alhag

## Abstract

**Background:** COVID-19 a pandemic declared by WHO, is the first in recent history pose challenges on public health. Health Belief Model is a psychosocial model explains and predicts health-related behaviours. This study aimed to explore the perceptions of the Sudanese on COVID-19-related preventive measures.

**Methods:** A Cross-sectional study using online-questionnaire was conducted between 1^st^-16^th^ April 2020 among Sudanese adults (aged ≥18 years). We used a snowball sampling technique, starting from known professional and social media groups, and individuals and then was distributed on various internet platforms. The survey instrument was based on HBM constructs.

**Results:** Some 877 individuals participated in the survey with a mean age 37.8 (SD±11.94) more males, mostly having a university education, employed and residing in Khartoum. More than half of the participants scored high in almost all Health Belief Model constructs, except for benefits of hand hygiene. The findings show that the HBM constructs are correlated to each other’s as well as to other socio-demographic factors. Self-efficacy correlated negatively with susceptibility (r −0.084), while positively with severity, benefits of and barriers to hand hygiene, benefits and barriers to social distancing (r 0.117, r 0.347, r 0.202, r 0.396, r 0.276), respectively.

**Conclusion:** The findings show that the HBM constructs are correlated to each other’s as well as to other socio-demographic factors. Self-efficacy must be taken into account as a strong changing factor to susceptibility and severity perceptions. Correlations found in this study might help drive behaviour-changing efforts.

## Background

Pneumonia of unknown cause detected in Wuhan; China was first reported to the World Health Organization (WHO) in China on 31 December 2019 (1). On 11 February 2020, WHO used the term 2019 novel coronavirus to refer to a coronavirus that affected the lower respiratory tract of the patients.

Based on further studies formally recognized this virus related to Severe Acute Respiratory Syndrome Coronavirus (SARS-CoV) and renamed it as SARS-CoV-2 (2). The coronavirus belongs to a family of viruses that are common in animals and may to affect humans. Cause various symptoms such as pneumonia, fever, breathing difficulty, and lung infection (2).

On 11 March, the coronavirus disease 2019 (COVID-19) outbreak spread to 46 countries and was declared by WHO as a pandemic, the first in recent history (3). The COVID-19 is highly transmissible as the effective reproductive number (R_0_) of COVID-19 (2.6-4.71) was higher than that reported of SARS (1.77) at this early stage (4-6). The average incubation duration of COVID-19 was estimated to be 4.8 days (SD± 2.6), ranging from (2 – 14 days). It tended to be shorter among people aged ≥70-years (7-9). The global mortality rate is about 5.7% (10). Early studies reported that the median age of patients to be 59-years, ranging from 15 to 89 years, with the majority (59%) being males (11, 12). Worldwide more than 4 million affected and more than and more than 300.000 deaths (13).

There are different sources from which the public receives the information about the COVID-19 cases, and mortality rate and accordingly may have different knowledge and different perceived severity and susceptibility to the disease. All of these could associate with the public’s emotional and behavioural reactions towards the COVID-19. The Public behaviours are essential for outbreak management, particularly during the early phase when no treatment or vaccination is available, and the only option is the engagement in precautionary behaviours, such as wearing masks, hand hygiene and social distancing (14). One of the significant emotional reactions during a pandemic is fear. Humans, like other animals, possess a set of defensive systems for combating ecological threats; strong fear appeals produce the most significant behaviour change only when people feel a sense of efficacy, whereas persuasive fear appeals with low-efficacy messages produce the highest levels of defensive responses (15).

Health Belief Model (HBM) HBM is one of the first theories of health behaviour, it was developed in the 1950s, it assumes that the assessments of health behaviour modification depend on the balance between the barriers to and benefits of action (16, 17). HBM is a social and psychological health behaviour change model, which was developed to explain and predict health-related behaviours, particularly concerning the uptake of health services (18). The theoretical constructs that constitute the HBM are broadly defined; furthermore, the HBM does not specify how constructs of the model interact with one another. Consequently, different operationalization of the theoretical constructs may not be strictly comparable across studies (19, 20)

The HBM is health behaviour focused, and there is evidence that the effectiveness of interventions to promote health behaviour change and improve health outcomes could depend on the use of models like the HBM (16).

Perceptions or beliefs about an outbreak may be necessary for determining compliance with health advice (21). The public may be more likely to comply with health recommendations if they believe that they are effective (22). Also, the perceived susceptibility, severity and high likelihood that they may be affected by the outbreak (23).

In a study in China, showed that being older, having good health, having higher education, perceiving the virus to be more severe, and having more knowledge were positively related to more social compliance (24). The social compliance, risk perception and severity are not consistent across demographic variations, social status and overtime. Indeed, previous studies have shown that perceptions and behaviours often change over time (25).

On 13 March 2020, Sudan reported its first COVID-19 in Khartoum, and the cases exceeds 2500 and more than 100 deaths, in spite of the total lockdown and other measures (26). Sudan faces immense crisis due the political unstability and economic impact of the pandemic and poor health system.

Implementation of public health measures should be based on the understanding of the public’s perceptions, beliefs, and attitude; therefore, this study aimed to explore the roles of perceived threat (perceived susceptibility and perceived severity), benefits, and barriers on the health preventive measures towards COVID-19 among Sudanese population. The survey results will be useful to assess the subsequent interventions and communication strategies as the epidemic progresses.

## Method

A Cross-sectional online survey was conducted between 1^st^-16^th^ April 2020 among Sudanese adults (aged ≥18 years). Following the announcement of first cases of COVID-19 in the country in March 2020, precautionary measures were taken to increase preventive measures (social distancing, non-essential contact with others, and all unnecessary travel). A snowball sampling technique was used, starting from known professional and social media groups and individuals. To ensure high coverage, all those contacted were requested to share the survey link and promote messages on their webpages, social media platforms or any channels, which they usually use to convey information to their contacts, with no restriction on their dissemination. The online survey link was distributed on various internet platforms, including WhatsApp (the most popular app in Sudan), Facebook and Twitter. The link (https://forms.gle/D3VhQgQVEirFH3gF6), opening standardized instructions about the purpose of the study and the procedure for completion of the survey. Individuals who were aged ≥18-year living in Sudan were eligible to participate. The survey could not be taken twice from the same electronic device. Participation was voluntary, and no incentive rewards were given. Anonymity was ensured as no identifiable information was collected. The survey could be completed in less than ten minutes. If a participant filled and submitted the form, it was considered as a consent to the participation. The questionnaire was constructed in English and administered in Arabic. The questionnaire was translated from English into Arabic and subsequently back-translated into English by experts in both languages. A pilot study testing the accuracy of translation and understanding of the questions was conducted before administration of the study among selected participants. This pilot was conducted, including 20 participants (male, female). Some minor adjustments of the survey instrument were performed before it was administered in the survey.

A timeline of two weeks was set, with two reminders and the link was closed after that. Eight hundred seventy-nine individuals participated in the survey.

This study obtained approval from the Research Ethics Committee at the University of Science and Technology, Omdurman, Sudan UST.

### Measures

The survey instrument was based on HBM constructs of self-efficacy, perceived susceptibility and severity to COVID-19 benefits from and barriers to the preventive measures (17), beside socio-demographic characteristics, health-related information and COVID-19 related-history.

Respondents completed subscales assessing the HBM constructs. All items were rated on five-point Likert’s scales (from strongly disagree to strongly agree) literature and were averaged to create HBM constructs.

*Two items measured perceived susceptibility* to COVID-19; “how likely you will be infected” “how likely your family will be infected”. Using a five-point scale (5= strongly agree-1= strongly disagree). A sum variable “Perceived susceptibility to COVID-19” (Cronbach’s α 0.80) was constructed from the two items and was dichotomized on the mean (6.66± SD 1.96) with values (0) low susceptibility (initial score <6.66); (1) high susceptibility (initial scores ≥6.66).

*Three items measured perceived severity* if infected with COVID-19 was measured by using three items; “the seriousness of symptoms caused by CoV-19”; “if I get COVID-19, it will affect how I feel in everyday life”; “if I get COVID-19, I will lose my life”. Using a five-point scale (5= strongly agree-1=strongly disagree). A sum variable “perceived severity if infected with COVID-19” (Cronbach’s α 0.68) was constructed from the three items and was dichotomized on the mean (11.9 ± SD 2.4) with values (0) low severity (initial score <11.9), (1) high severity (initial scores >11.9).

*Two items measured barriers to benefits of hand hygiene*; “hand hygiene will always prevent me from getting COVID-19”, “I feel safer through hand hygiene”. Using a 5-point scale (5= strongly agree-1=strongly disagree). Using a five-point scale (5= strongly agree-1=strongly disagree). A sum variable “benefits of hand hygiene to prevent COVID-19” (Cronbach’s α 0.68) was constructed from the two items and was dichotomized on the mean (8.18 ± SD 1.76) with values (0) no (initial score <8.18); (1) yes (initial scores ≥8.18).

*Two items measured barriers to hand hygiene*; “my hands hurt when I do hand hygiene”; “I always forget hand hygiene”. Using a five-point scale (5= strongly agree-1=strongly disagree). A sum variable “barriers of hand hygiene to prevent COVID-19” (Cronbach’s α 0.10) was constructed from the two items and was dichotomized on the mean (7.20 SD ±1.62) with values (0) no (initial score <7.20); (1) Yes (initial scores ≥7.20).

*Two items measured benefits of social distancing*; “the social distancing prevents me from getting COVID-19”; “I feel safer by social distancing”. Using five-point scale (5 = strongly agree-1=strongly disagree). A sum variable “benefits of social distancing to prevent COVID-19” (Cronbach’s α 0. 83) was constructed from the two items and was dichotomized on the mean (9.07± SD 1.18) with values (0) no (initial score <9.07); (1) yes (initial scores ≥9.07).

*Two items measured barriers to social distancing*; “I feel bad with social distancing”; “I always forget social distancing”. Using a five-point scale (5= strongly agree-1=strongly disagree). A sum variable “barriers of social distancing to prevent COVID-19” (Cronbach’s α 0. 48) was constructed from the two items and was dichotomized on the mean (7.22± SD 1.87) with values (0) no (initial score <7.22); (1) yes (initial scores ≥7.22).

*Five items measured self-efficacy* towards health COVID-19; “maintaining good health is an important part of my life” and “I think I am a person who cares well for his general health”.” I think it is important for me to have good general health”, “I think it is important for me to avoid infectious diseases” and “I think I am a person who takes correct health measures”. Using a five-point scale (5= strongly agree-1=strongly disagree). A sum variable “self-efficacy to prevent COVID-19” (Cronbach’s α 0. 73) was constructed from the two items and was dichotomized on the mean (22.44±2.21 SD) with values (0) no efficacy (initial score <22.44); (1) yes (initial scores ≥22.44).

### Data analysis

We analyzed data using the Statistical Package for the Social Science, version 23 (IBM SPSS). We performed descriptive analyses using frequencies and percentages for categorical variables, mean and Standard Deviation (±SD) for numerical variables. Bivariate relationships between the dependent variables and independent variables were assessed using t-test. We conducted multiple variable analysis using the correlation between HBM constructs. Estimates are presented as Pearson Correlation coefficients (r) with 95% confidence interval (CI), a two-sided significance level of ≤ 5% was implied for all analyses.

## Results

### Sample profile

A number of 877 individuals participated in the survey with a mean age 37.8 (SD±11.94), most of the participants (77%) were in the age-group 24-55-year-old. Males were slightly higher than the females, 59% (517). The majority had university or higher education 94.4% (828), most of them 73% (636) were employed, and resided in Khartoum State 76.4% (670), with almost all had no history of travelling during the past 14 days 92% (807) (Figure 1).

**Figure (1):**
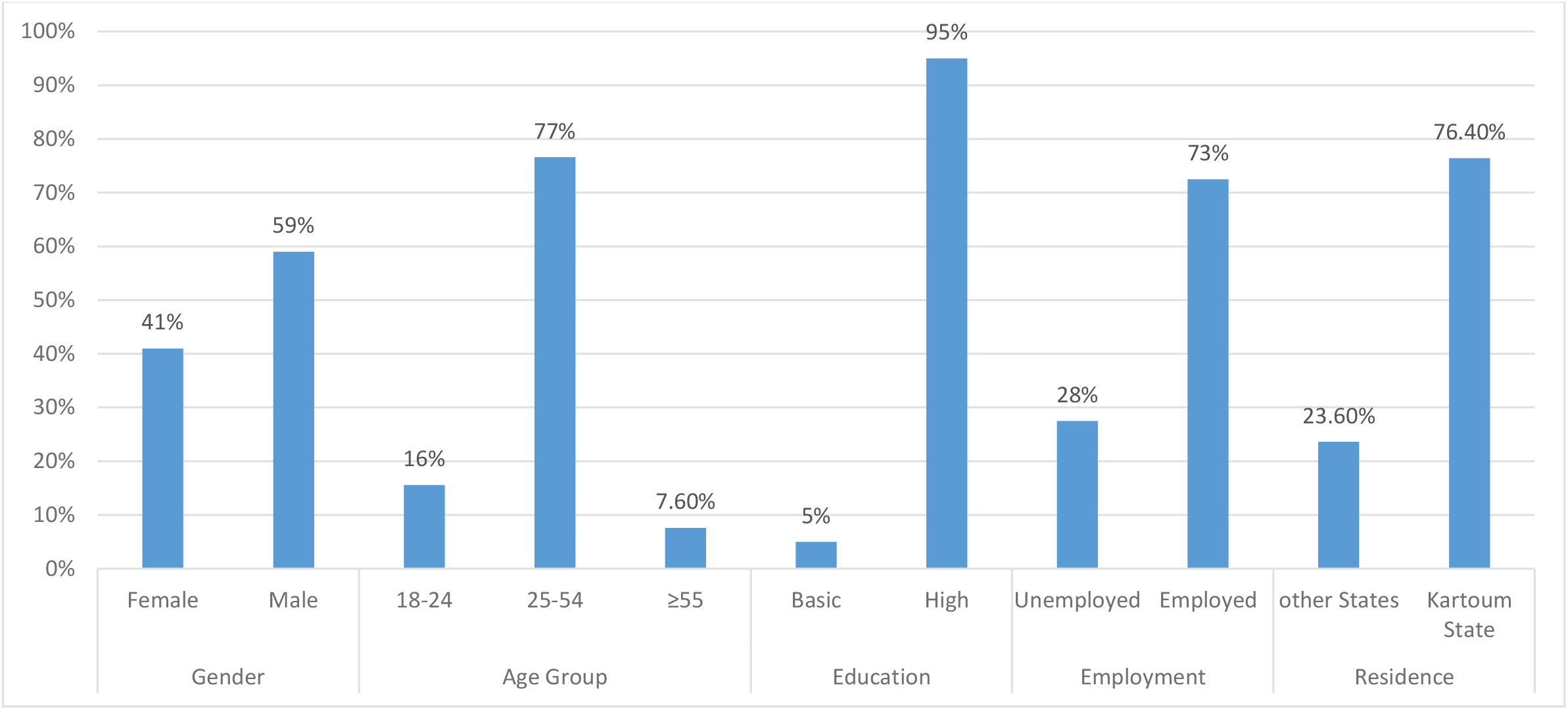
Percentages (%) distribution of the socio-demographic characteristics of the participants.

Regarding reported health status, most of the participants perceived having good/very good health status 87.8% (770), reported no medical consultation in the past month 86.2% (756), no respiratory symptoms 74.2% (651), and no organic/mental illness 75.3% (661). Almost all the participants reported no relation neither with COVID-19 infection nor with anyone with COVID-19 infection 97.9% (859), 88.9% (867) (Figure 2).

**Figure (2):**
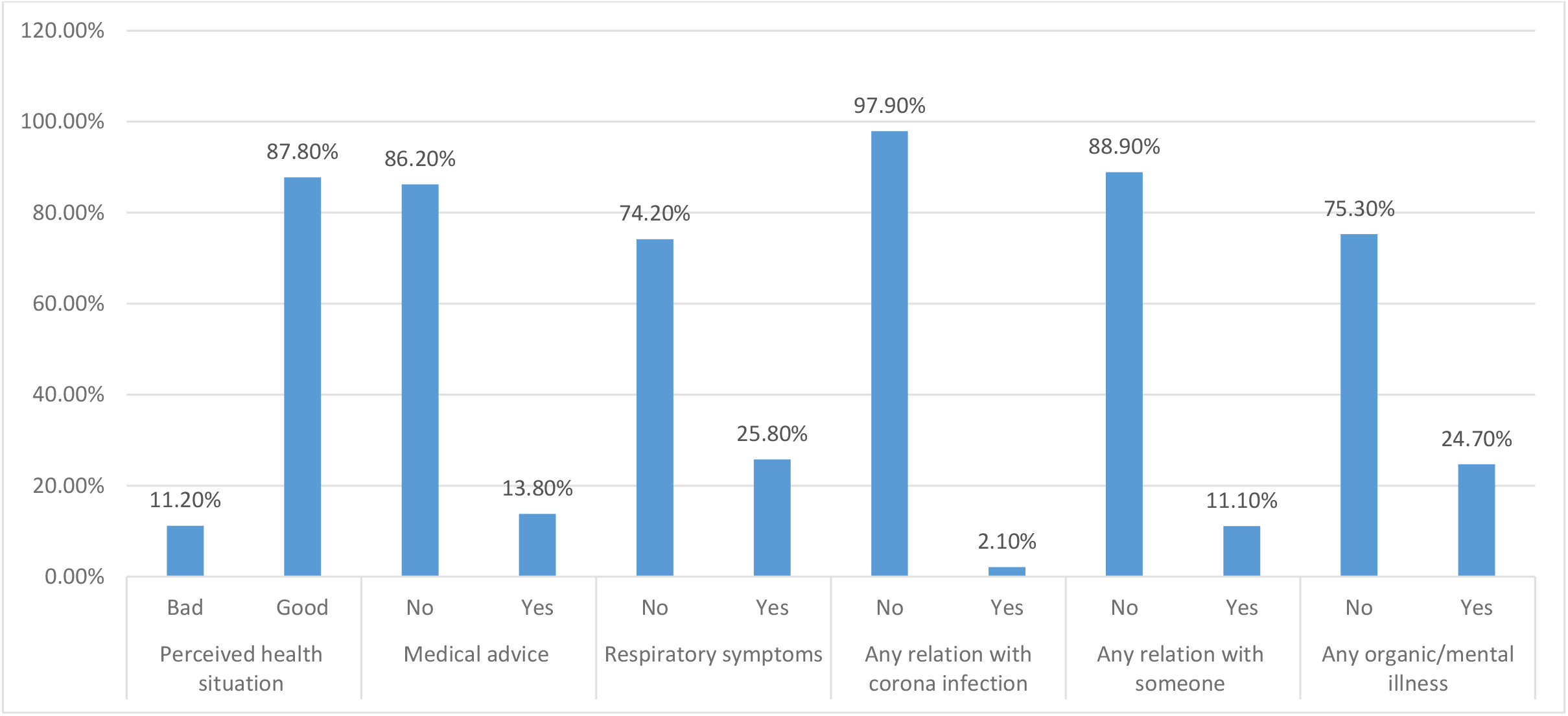
Percentages distribution of reported health-related aspects.

### Health Belief Model Constructs

Looking at Self-efficacy individual statements, almost all the participants agreed/strongly agreed to the five statements. Regarding susceptibility statements, most of the participants either agreed or were neutral to both statements. Majority of the participants agreed/strongly agreed to the three statements of the severity construct, as well as the benefits of hands hygiene. About statements of the barriers to hands hygiene, participants varied with almost half agreed to the first while disagreed to the second statement (Table 1).

**Table 1:**
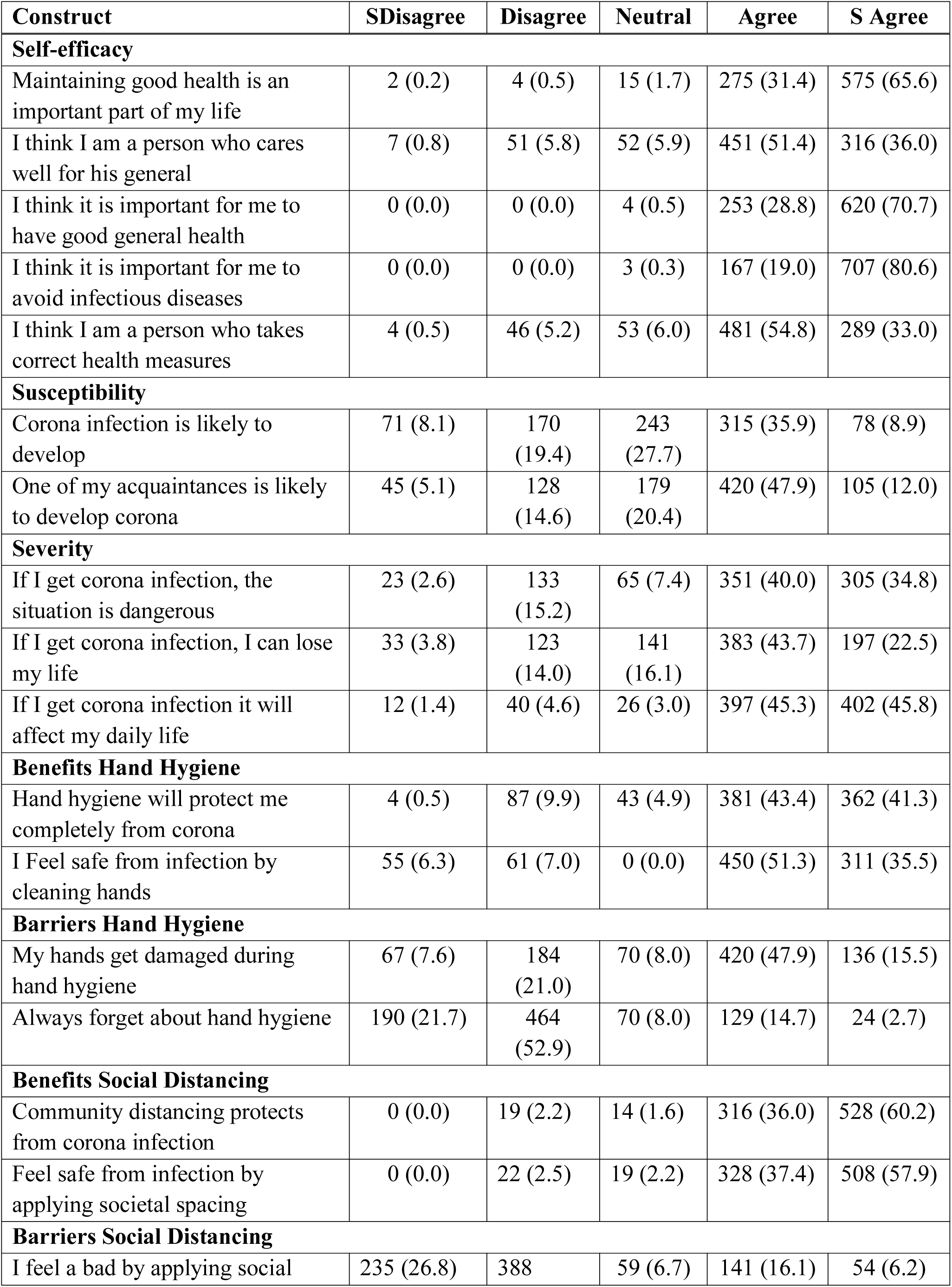
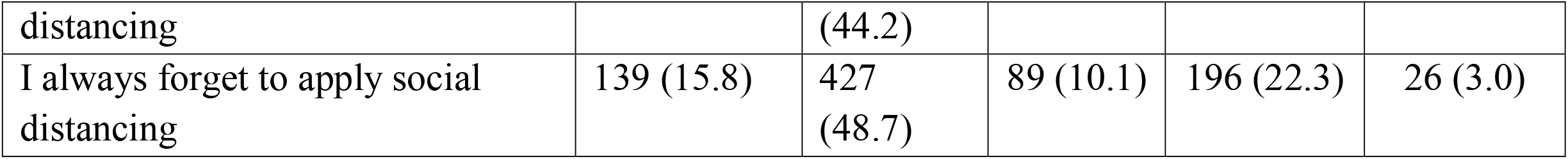
Frequencies (n) and percentages (%) of Health belief model constructs.

When HBM constructs were dichotomized, the participants’ scores were divided almost equally through all the constructs after being dichotomized using the mean scores. Participants scored high in almost all constructs ranging from 52% to 60%, except for barriers and benefits hand hygiene. The participants were almost equally distributed 50.3% versus 49.7% regarding barriers hand hygiene, but benefits hand hygiene was the single constructs where more participants scored low benefits 54% versus 46% (Figure 3).

**Figure (3):**
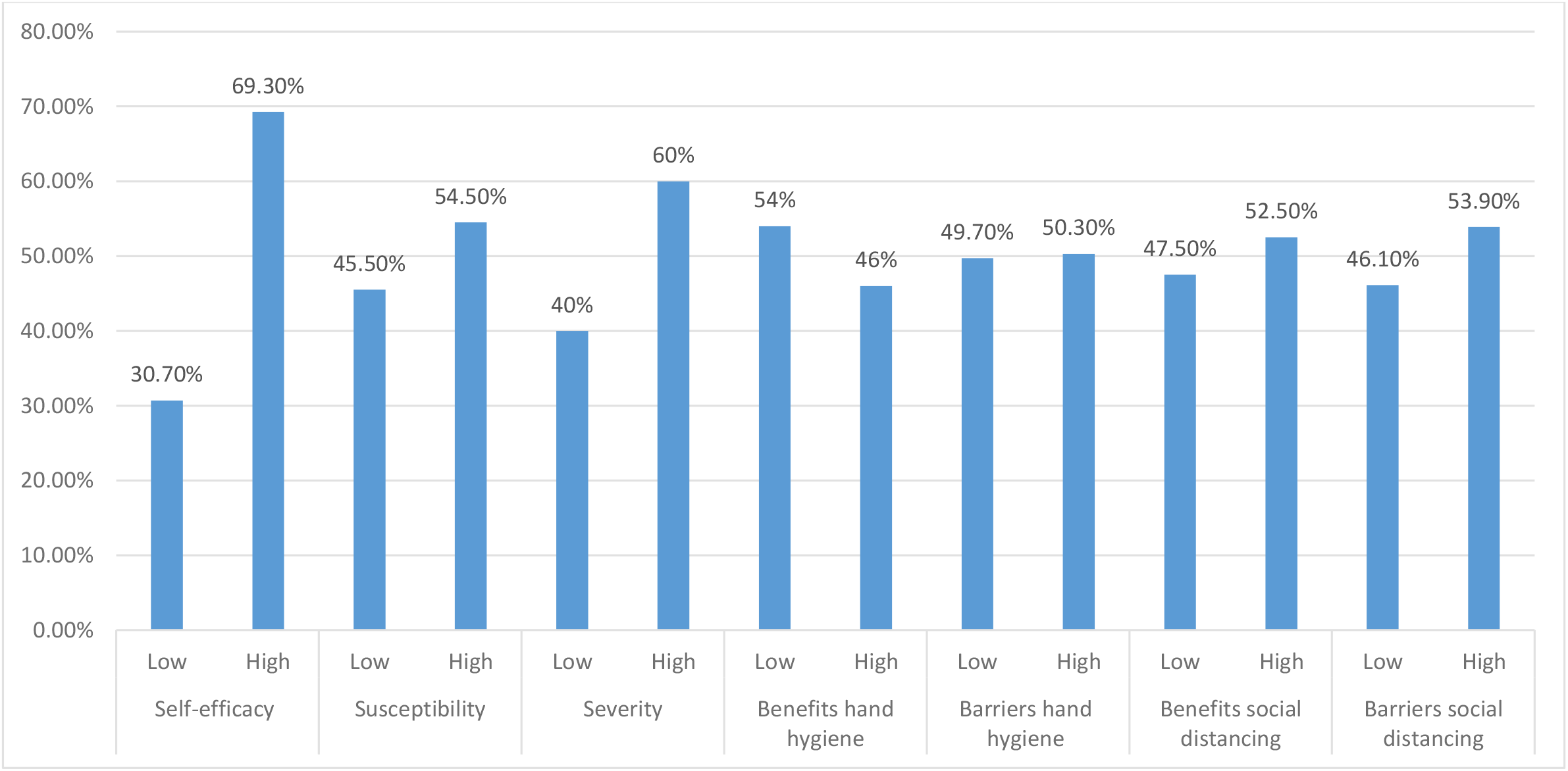
Percentage distribution of HBM Constructs.

There were statistically significant differences among gender, as males perceived higher barriers to hand hygiene, while females perceived higher benefits of and barriers to social distancing. Many statistically significant differences were observed: Those who reported a lower level of education perceived higher benefits hand hygiene. Participants with a history of medical consultation last 14 days perceived higher susceptibility compared to their counterparts. Those with respiratory symptoms perceived higher susceptibility to and severity of the infection and threat perception; they perceived fewer barriers to hands hygiene and less self-efficacy (Table 2).

**Table 2:** Mean differences SE, and CI of HBM Constructs by personal and health-related aspects.

| **Variable** | **Mean** | **M difference** | **SE** | **CI** | **P-value** |
| --- | --- | --- | --- | --- | --- |
| **Gender** (Female/Male) |  |  |  |  |  |
| Barriers hand hygiene | 7.01/7.30 | -0.28 | 0.11 | -0.50, -0.07 | 0.011 |
| Benefits social distancing | 9.17/8.96 | 0.21 | 0.08 | 0.05, 0.37 | 0.010 |
| Barriers social distancing | 7.47/7.30 | 47.0 | 0.13 | 0.19, 0.68 | 0.001 |
| **Education** Low/High |  |  |  |  |  |
| Benefits hand hygiene | 8.70/8.14 | 0.55 | 0.20 | 0.15, 0.96 | 0.008 |
| **Medical advice** (No/Yes) |  |  |  |  |  |
| Susceptibility | 6.6/7.0 | -0.43 | 0.19 | -0.81, -0.06 | 0.024 |
| **Respiratory symptoms** (No/Yes) |  |  |  |  |  |
| Susceptibility | 6.5/7.0 | -0.50 | 0.14 | -0.78, -0.23 | 0.000 |
| Severity | 15.6/16.2 | -0.62 | 0.17 | -0.95, -0.28 | 0.000 |
| Barriers hand hygiene | 7.27/6.96 | 0.30 | 0.12 | 0.06, 0.55 | 0.013 |
| Self-efficacy | 22.6/21.9 | 0.62 | 0.18 | 0.28, 0.97 | 0.000 |
| Threat (Risk) perception | 22.1/23.2 | -1.12 | 0.25 | -1.6, 1.73 | 0.000 |
| **Travel outside Sudan past month**<br>(No/Yes) | 15.77/15.05 | 0.72 | 0.30 | 0.12, 1.32 | 0.019 |
| Severity | 22.4/21.5 | 0.93 | 0.041 | 0.13, 1.7 | 0.023 |
| Threat (Risk) perception |  |  |  |  |  |
| **Any relation with corona infection** (No/Yes) | 6.62/8.0 | -1.38 | 0.25 | -1.90, -0.85 | 0.000 |
| Susceptibility | 15.69/17.16 | -1.47 | 0.49 | -2.51, -0.44 | 0.008 |
| Severity | 22.3/25.2 | -2.85 | 0.8 | -3.8, -1.8 | 0.000 |
| Threat (Risk) perception |  |  |  |  |  |
| **Any relation with someone with corona** (No/Yes) | 15.71/16.70 | -0.99 | 0.37 | -1.83, -.15 | 0.025 |
| Severity | 9.04/9.7 | -0.66 | 0.22 | -1.14, -0.17 | 0.013 |
| Benefits social distancing |  |  |  |  |  |
| **Any organic or mental illness** (No/Yes) | 6.54/6.96 | -0.41 | 0.14 | -0.68, -0.15 | 0.003 |
| Susceptibility | 8.25/7.95 | 0.30 | 0.14 | 0.02, 0.58 | 0.037 |
| Benefits hand hygiene | 22.2/22.7 | -0.50 | 0.24 | -.97, -.04 | 0.035 |
| Threat (Risk) perception |  |  |  |  |  |

Participants who had travelling history in the last month perceived less severity and threat perception of the infection compared to their counterparts. Participants reporting no any relation with the infection perceived less susceptibility and severity to the infection, and those reporting no relation with anyone affected with the infection scored less severity and benefits of social distancing (Table 3, 4).

**Table 3:** Mean (±SD), Min-Max, and CI of HBM constructs.

| Variable | Mean (±SD) | SD | CI | Min/ Max |
| --- | --- | --- | --- | --- |
| Self-efficacy | 22.44 | 2.21 | 22.29, 22.59 | 14/25 |
| Susceptibility | 6.66 (1.96) | 2.0 | 6.53, 6.79 | 2/10 |
| Severity | 11.9 (2.4) | 2.3 | 11. 7, 12.0 | 3/15 |
| Benefits hand hygiene | 8.18 | 1.76 | 8.07, 8.30 | 2/10 |
| Barriers hand hygiene | 7.20 | 1.62 | 7.09, 7.30 | 2/10 |
| Benefits social distancing | 9.07 | 1.18 | 8.99, 9.15 | 4/10 |
| Barriers social distancing | 7.22 | 1.87 | 7.10, 7.35 | 2/10 |
| Threat | 22.4 | 3. 2 | 22.2, 22.6 | 10/30 |

**Table 4:**
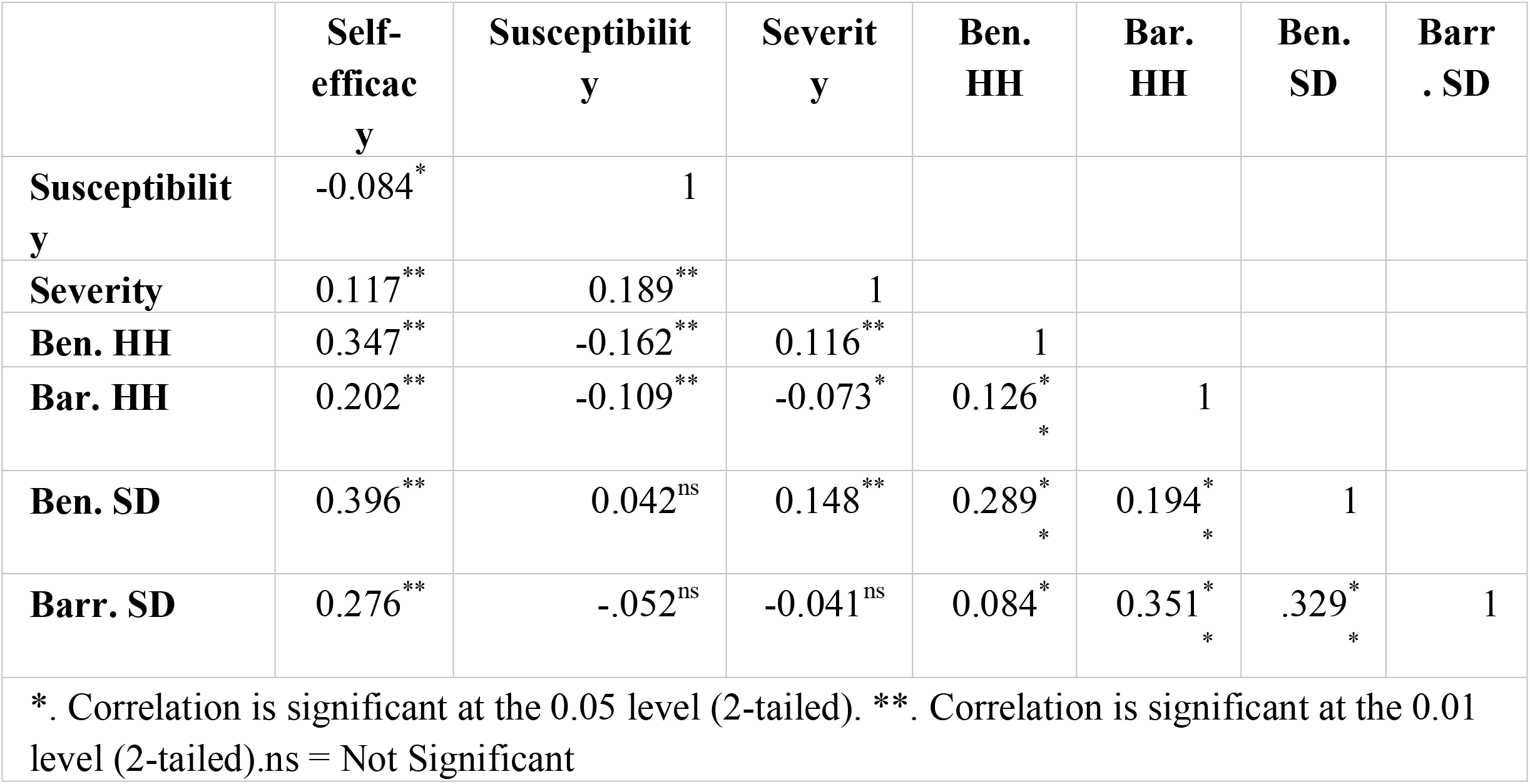
Correlation of the HBM constructs.

When HBM constructs were regressed, almost all constructs correlated statistically significant to each other except for susceptibility to benefits and barriers to social distancing, as well as the severity of barriers to social distancing.

Self-efficacy correlated positively with all the constructs and negatively with susceptibility. The correlation was stronger with benefits social distancing followed by benefits hands hygiene, followed by barriers to social distancing and barriers hands hygiene. The least correlated were severity and susceptibility.

On the other hand, susceptibility negatively correlated with self-efficacy, barriers, and benefits hand hygiene, while correlated positively with severity, which was the strongest correlation, followed by benefits hands hygiene. The least correlation was with severity.

Severity correlated with self-efficacy, susceptibility, benefits hand hygiene and social distancing, while correlated negatively with barriers to hands hygiene.

Benefits hand hygiene correlated positively with self-efficacy, severity, benefits and barriers social distancing, while correlated negatively only with susceptibility. Barriers hands hygiene correlated positively with self-efficacy benefits hand hygiene, benefits and barriers social distancing, while correlated negatively with susceptibility and severity.

Benefits social distancing correlated positively with all other constructs. Barriers social distancing correlated positively with all constructs except susceptibility and severity. Threat (risk) perception construct correlated positively with susceptibility severity and benefits social distancing, while negatively with barriers to hand hygiene (Table 4).

## Discussion

Application of psychological theories would provide systematic explanations about observable facts. Health behavior research has explored the effectiveness and applicability of various health models in health behavior modification (27).

According to the framework of the HBM in the context of COVID-19. Suggests that a person would be more likely to comply with recommended preventive behaviours if he/she perceives being susceptible to the infection (perceived susceptibility), and that the infection could lead to serious consequences (perceived severity).

Perceived barriers ((beliefs about obstacles to performing a behavior, and the negative aspects of adopting a health behavior) of social distancing and hand hygiene), and perceived benefits (belief in efficacy of the advised action to reduce risk or seriousness of impact) (28), are also important factors in taking actions on preventive measures. Self-efficacy has been shown to relate to various health-related practices, such as smoking cessation, diet, and health-promoting lifestyle (29, 30).

This study provides a timely assessment of the perceived self-efficacy, severity, susceptibility to COVID-19. Besides, exploring the barriers and benefits of the preventive measures (hand hygiene and social distancing) recommended by health authorities in Sudan using HBM.

The sample is to be considered realistic although less than expected for several reasons; Sudan literacy constitutes 75.9%, and the total internet users are only 26.4 % of the population in which only 8.3% is the active users of social media platforms where the online survey posted (31). The gender distribution among the participants almost similar to the population as the ratio of males to females (1.44-1.01) respectively.

Self-efficacy referring to the confidence in one’s ability to take action and successfully execute preventive health behavior (28). Among the participants, almost one-third of them scored low perceived self-efficacy. That means lack the self-efficacy to follow the guidelines of the preventive measures, which includes hand hygiene practices and social distancing to slow the spread of SARS/CORONA-2 virus (32). Perceived high self-efficacy is inversely related to perceived susceptibility, and positively related to perceived severity leading to more people adopting preventive measures, and is consistent with other studies (14, 24).

The previous experiences and literature, indicate that an emphasis on sustained interventions towards changing social norms yields the most effective results (33). Mitigating behaviours require significant efforts to strengthen beliefs about disease, which includes self-efficacy. Self-efficacy has been increasingly associated with health behaviour change, and it is a strong predictor of health-promoting behaviours (34, 35).

The higher the levels of perceived barriers of social distancing and hand hygiene, the more likely participants were to perceive high self-efficacy, and this is not in accordance to other studies in which negative relation where established between these constructs (36, 37).

Males perceived higher barriers to hand hygiene and social distancing than females. This is in line with the literature that females perceived higher benefits (38). Females’ prompted compliance of preventive measures in protecting themselves and others related to health issues. Perhaps targeting health promotion messages through females (for example, mothers, wives) who are more health-conscious and risk-averse would be worth exploring in an attempt to raise the level of protective precautions undertaken by this vulnerable subgroup (39, 40).

This study reported that the observed low perceived susceptibility (beliefs about the likelihood of getting a disease or condition) and severity (beliefs about the seriousness of contracting a disease or condition, including consequences) 45%-40% respectively (28), and this contradicts similar study in Hong Kong in which high perceived susceptibility and severity were reported (89%-97%) (14). Respondents who had respiratory, health-related issues, or relation with coronavirus perceived higher susceptibility, severity and threat of the infection, which is consistent with other responses during other epidemics like SARS (39).

This wide gap may be explained by as in China where the disease started far distance from Sudan, and here comes the role of “optimism bias” which is the belief that bad things are less likely to happen to oneself than others (41).

While optimism bias may be beneficial for avoiding anxiety emotions during a pandemic, it can lead people to underestimate their chance of contracting a disease, and therefore disregard public health warnings (42).

Another reason is fear known as a factor that can lead to behavioural changes among people. Fear was the most significant change in behaviours among those perceived high self-efficacy and produced defensive responses among those who perceived low efficacy towards their health (43). Hence, communication strategies in Sudan should follow a balance between breaking through optimism bias without inducing excessive fear

### Strengths and limitations

An apparent strength of this study is that data collection took place during the COVID-19 pandemic, where the threat was regarded as high, and the scenarios were not based on hypothetical situations. Another strength is that this study is based on HBM, enabling analysis and comparison with other studies using the model to explain health behaviour. Although these findings give valuable insight into health behaviours among the Sudanese population, several limitations should be noted. Using a web-based questionnaire might lead to selection bias. The only internet users who responded to our online questionnaire were not fully representative of the general population. Therefore, findings of this study should be cautiously interpreted and generalized. Considering use of self-report questionnaires, cross-sectional design, low response rate and small sample size.

### Conclusions

The findings show that the HBM constructs are correlated to each other’s as well as to other socio-demographic factors. Self-efficacy must be taken into account as a strong changing factor to susceptibility and severity perceptions. Correlations found in this study might be helpful in driving behavior –changing efforts.

## Data Availability

All data will be available upon request

## Contribution Statement

The roles of the authors were; EFN, HMA, AKA contributed to the conception and design of the study, the acquisition of data, analysis and interpretation of data. All authors contribute to write the paper, have critically read and edited the paper. All authors have read and approved the final manuscript.

## Acknowledgement

None

## References

1. WHO. Novel Coronavirus–China. 2020. Available on: https://www.who.int/csr/don/12-january-2020-novel-coronavirus-china/en/. Accessed 17 May 2020 [

2. Chen Y, Liu Q, Guo D. Emerging coronaviruses: Genome structure, replication, and pathogenesis. J Med Virol. 2020;92(4):418–23. doi: 10.1002/jmv.25681.

3. WHO. WHO Director-General’s opening remarks at the media briefing on COVID-19 - 11 March 2020. Available on: https://www.who.int/dg/speeches/detail/who-director-general-s-opening-remarks-at-the-media-briefing-on-covid-19---11-march-2020. Accessed 17 May 2020 [

4. Liu T, Hu J, Kang M, Lin L, Zhong H, Xiao J, et al. Time-varying transmission dynamics of Novel Coronavirus Pneumonia in China. bioRxiv 2020 0125919787; doi: https://doiorg/101101/20200125919787.

5. Adhikari SP, Meng S, Wu Y-J, Mao Y-P, Ye R-X, Wang Q-Z, et al. Epidemiology, causes, clinical manifestation and diagnosis, prevention and control of coronavirus disease (COVID-19) during the early outbreak period: a scoping review. Infect Dis Poverty. 2020 Mar 17;9(1):29. doi.org/10.1186/s40249-020-00646-x.

6. Liu Y, Gayle AA, Wilder-Smith A, Rocklöv J. The reproductive number of COVID-19 is higher compared to SARS coronavirus. J Travel Med. 2020 Mar 13; 27(2):taaa021.doi:10.1093/jtm/taaa021.

7. Wang W, Tang J, Wei F. Updated understanding of the outbreak of 2019 novel coronavirus (2019-nCoV) in Wuhan, China.. J Med Virol. 2020;92(4):441–7.doi:10.1002/jmv.25689.

8. Lauer SA, Grantz KH, Bi Q, Jones FK, Zheng Q, Meredith HR, et al. The Incubation Period of Coronavirus Disease 2019 (COVID-19) From Publicly Reported Confirmed Cases: Estimation and Application.. Ann Intern Med. 2020;172(9):577–82.doi:10.7326/M20-0504.

9. National Health Commission of People’s Republic of China. New coronavirus infection pneumonia public protection guidelines. Available on: http://www.nhc.gov.cn/xcs/yqfkdt/202001/bc661e49b5bc487dba182f5c49ac445b.shtml. Accessed 17 May 2020. [

10. Baud D, Qi X, Nielsen-Saines K, Musso D, Pomar L, Favre G. Baud D, Qi X, Nielsen-Saines K, Musso D, Pomar L, Favre G. Real estimates of mortality following COVID-19 infection [published online ahead of print,. Lancet Infect Dis. 2020 Mar 12];S1473–3099(20):30195-X.doi:10.1016/S1473-3099(20)-X.

11. Li Q, Guan X, Wu P, Wang X, Zhou L, Tong Y, et al. Early Transmission Dynamics in Wuhan, China, of Novel Coronavirus-Infected Pneumonia. N Engl J Med. 2020;382(13):1199–207.doi:10.056/NEJMoa2001316.

12. Chinese Center for Disease Control and Prevention. The Epidemiological Characteristics of an Outbreak of 2019 Novel Coronavirus Diseases (COVID-19) — China, 2020 The Novel Coronavirus Pneumonia Emergency Response Epidemiology Team. avialable on: https://cdn.onb.it/2020/03/COVID-19.pdf. Accessed 17 May 2020. CCDC Weekly / Vol 2 / No 8. 2020;2(8):1–10.

13. JOHNS HOPKINS UNIVERSITY AND MEDICINCE, Coronavirusresource Center. Available on: https://coronavirus.jhu.edu/?utm_source=jhu_properties&utm_medium=dig_link&utm_content=ow_jhuhomepage&utm_campaign=jh20 Accessed 17 May 2020.

14. Kwok KO, Li KK, Chan HH, Yi YY, Tang A, Wei WI, et al. Community Responses during Early Phase of COVID-19 Epidemic, Hong Kong [published online ahead of print, 2020 Apr 16]. Emerg Infect Dis. 2020;26(7):10.3201/eid2607.200500. doi:10.3201/eid2607.

15. Witte K, Allen M. A meta-analysis of fear appeals: implications for effective public health campaigns.. Health Educ Behav. 2000;27(5):591–615.doi:10.1177/109019810002700506.

16. Munro S, Lewin S, Swart T, Volmink J. A review of health behaviour theories: how useful are these for developing interventions to promote long-term medication adherence for TB and HIV/AIDS? BMC public health. 2007 Jun 11;7:104.doi:10.1186/471-2458-7-104.

17. Hochbaum G. Health Behavior: Belmont, CA: Wadsworth Publishing; 1970

18. Maiman LA, Becker MH, Kirscht JP, Haefner DP, Drachman RH. Scales for Measuring Health Belief Model Dimensions: A Test of Predictive Value, Internal Consistency, and Relationships among Beliefs. Health Education & Behavior. 1977;5(3):215–31.

19. Glanz K, Bishop DB. The role of behavioral science theory in development and implementation of public health interventions.. Annu Rev Public Health. 2010;31:399–418.doi:10.1146/annurev.publhealth.012809.103604.

20. Orji R, Vassileva J, Mandryk R. Towards an effective health interventions design: an extension of the health belief model.. Online J Public Health Inform. 2012;4(3):ojphi.v4i3.4321.doi:10.5210/ojphi.v4i3.4321.

21. Rubin GJ, Amlôt R, Page L, Wessely S. Public perceptions, anxiety, and behaviour change in relation to the swine flu outbreak: cross sectional telephone survey.. Bmj. 2009 Jul 2;339:b2651.doi:10.1136/bmj.b2651.

22. Lau J, Yang X, Tsui H, Kim J. Monitoring community responses to the SARS epidemic in Hong Kong: from day 10 to day 62.. J Epidemiol Community Health. 2003;57(11):864–70.doi:10.1136/jech.57.11.864.

23. Tang CS-k, Wong C-y. Factors influencing the wearing of facemasks to prevent the severe acute respiratory syndrome among adult Chinese in Hong Kong.. Prev Med. 2004;39(6):1187–93.doi:10.016/j.ypmed.2004.04.032.

24. Li J-B, Yang A, Dou K, Wang L-X, Zhang M-C, Lin X. Chinese public’s knowledge, perceived severity, and perceived controllability of the COVID-19 and their associations with emotional and behavioural reactions, social participation, and precautionary behaviour: A national survey. Research Square. 2020.

25. Atchison CJ, Bowman L, Vrinten C, Redd R, Pristera P, Eaton JW, et al. Perceptions and behavioural responses of the general public during the COVID-19 pandemic: A cross-sectional survey of UK Adults. doi - 10.1101/2020.04.01.20050039. medRxiv. 2020.

26. Sudan Federal Ministry Of Health. Available on: https://www.facebook.com/FMOH.SUDAN/. Accessed 19 May 2020.

27. Hollister MC, Anema MG. Health behavior models and oral health: a review. J Dent Hyg. 2004;78(3):6.

28. Glanz K, Rimer BK, Viswanath K. Health Behavior and Health Educatheory, Research, and Practice. pp 48. 4th ed: ohn Wiley & Sons; 2015.

29. Berner ES. Clinical decision support systems. second ed: Springer; 2007.

30. Hollister MC, Anema MG. Health behavior models and oral health: a review. J Dent Hyg. 2004;78(3):6.

31. Internet live stats. Sudan Inernet Users. available at https://www.internetlivestats.com/internet-users/sudan/. Accessed on 17 May 2020. [

32. WHO. Coronavirus disease (COVID-19) advice for the public. Avialable on : https://www.who.int/emergencies/diseases/novel-coronavirus-2019/advice-for-public.. Accessed 17 May 2020. [

33. Merzel C, D’Afflitti J. Reconsidering community-based health promotion: promise, performance, and potential. Am J Public Health. 2003;93(4):557–74.doi:10.2105/ajph.93.4.557.

34. Bandura A. Self-efficacy: toward a unifying theory of behavioral change.. Psychol Rev. 1977;84(2):191–215.doi:10.1037//0033-295x.84.2.191.

35. Mukhtar S. Mental health and emotional impact of COVID-19: Applying Health Belief Model for medical staff to general public of Pakistan Brain Behav Immun. 2020 Apr 10;S0889–1591(20):30463-3.doi:10.1016/j.bbi.2020.04.012.

36. Anagnostopoulos F, Buchanan H, Frousiounioti S, Niakas D, Potamianos G. Self-efficacy and oral hygiene beliefs about toothbrushing in dental patients: a model-guided study.. Behav Med. 2011;37(4):132–9.doi:10.1080/08964289.2011.636770.

37. Baghiani Moghadam M, Mozaffarian Azad M, Biria M, Sabour S. Evaluation of oral hygiene care of under 4 years old children by their mothers based on the Health Belief Model. J Dent Sch 2015;33(1):9–18.

38. Zetu L, Zetu I, Dogaru CB, Duta C, Dumitrescu AL. Gender variations in the psychological factors as defined by the extended health belief model of oral hygiene behaviors. Social and Behavioral Sciences 2014;127:358–62.doi: 10.1016/j.sbspro.2014.03.271

39. Leung G, Lam T, Ho L, Ho S, Chan B, Wong I, et al. The impact of community psychological responses on outbreak control for severe acute respiratory syndrome in Hong Kong. J Epidemiol Community Health. 2003;57:857–63.

40. Bults M, Beaujean DJ, de Zwart O, Kok G, van Empelen P, van Steenbergen JE, et al. Perceived risk, anxiety, and behavioural responses of the general public during the early phase of the Influenza A (H1N1) pandemic in the Netherlands: results of three consecutive online surveys. BMC public health. 2011 Jan 3;11:2. doi:10.1186/471-2458-11-2.

41. Sharot T. The optimism bias. Curr Biol. 2011;21(23):R941–R5.doi:10.1016/j.cub.2011.10.030.

42. Van Bavel JJ, Boggio P, Capraro V, Cichocka A, Cikara M, Crockett M, et al. Using social and behavioural science to support COVID-19 pandemic response Nat Hum Behav. 2020 Apr 30;10.1038:s41562-020-0884-z.doi: 10.1038/s-020-0884-z.

43. Witte K, Allen M. A meta-analysis of fear appeals: Implications for effective public health campaigns. Health education & behavior. 2000;27(5):591–615.

